## Supporting Information for "Future prevalence of type 2 diabetes – a comparative analysis of chronic disease projection methods"

### Comparison of prevalence projections assuming constant prevalence vs. using method 2) or 3)

S1 Fig shows the age-specific prevalence for males in Germany from 2010 to 2040 assuming rising general life expectancy, i.e., variant B1L2M1 of the FSO population projection. Method 1) assumes that the prevalence remains constant, while method 2) reflects on temporal trends affecting the prevalence. Method 3) does not require an intermediate step to compute the prevalence first, but instead directly returns projected case numbers. Though, implicitly, we can calculate the prevalence with method 3) using the proportion of $I(t,\ a)$ and the sum of $I(t,\ a)$ and $H(t,\ a)$, i.e. $p(t,a) = \frac{I(t,a)}{I(t,a)+H(t,a)}$.

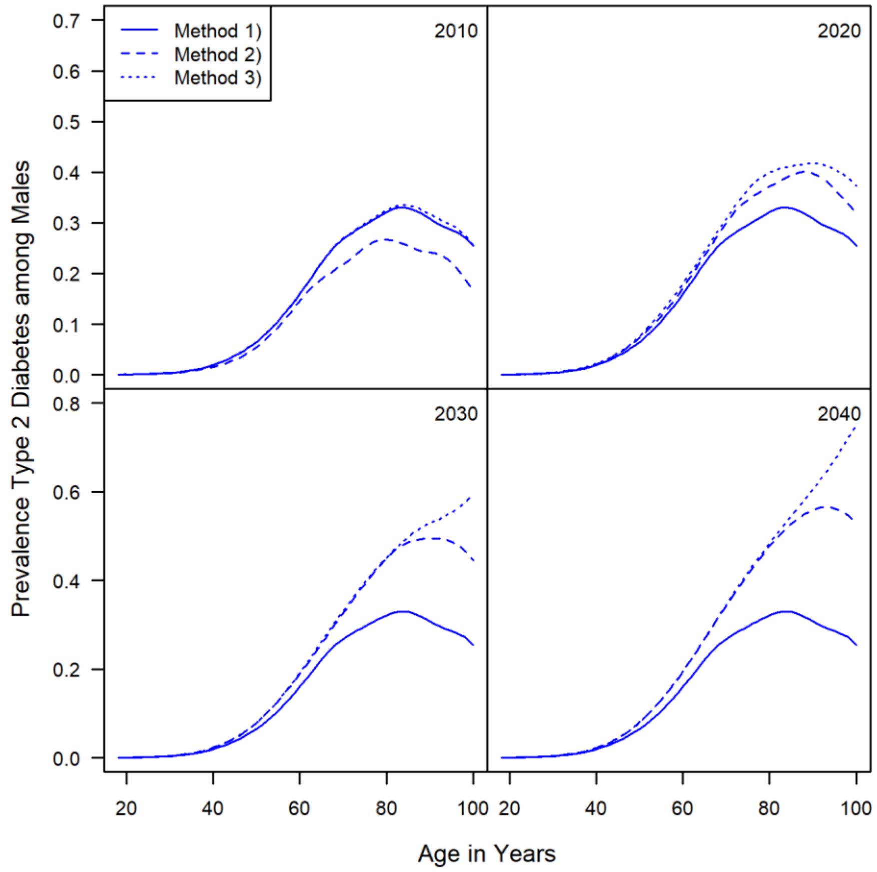


***S1 Fig. Comparing T2D prevalence projections.*** *Projected age-specific prevalence of T2D among males in Germany in 2015, 2020, 2030 and 2040 for method 1), 2) and 3). Projections are based on variant B1L2M1 of the FSO population projections. Method 1) assumes constant age-specific prevalence, method 2) and 3) assume a constant T2D incidence rate but a yearly decrease of 2% in the MRR.*

The figure shows apparent discrepancies in the estimation of the age-specific prevalence among method 1) and the other two methods. Compared to assuming a constant T2D prevalence as in method 1), the prevalence computed with method 2) and 3) is expected to increase markedly, especially for all ages above 60 years. Aside from that, the peak prevalence is presumed to shift towards older age groups. In 2015, the prevalence reaches its highest point of 30-40% at approximately 80 years of age, while in 2040 the prevalence peaks at about 90 years of age, lying between 50-60%. The smallest increases are expected in the younger age groups below 60 years of age.

The projected prevalence of method 2) and 3) resemble each other in terms of the course of the estimated lines as well as in the intensity. Though, as of 2030, the strong increase in the estimated future prevalence for males older than 85 years using method 3) seems implausible. There are two possible explanations. Firstly, and as is visible for method 2), the peak prevalence generally shifts towards older age groups. Secondly, this is probably an artifact of the population pyramid in 2010 [18] which is used to calibrate the mortality rates $m_0$ and $m_1$ with method 3). The population data in 2010 is still marked by the noticeable increase in the birth rate during the so-called post–World War II baby boom. The generation of the baby boomers, i.e., people born around 1950, will belong to the critical age groups of 85 years and above in year 2030 which is then mirrored in the high numbers of projected T2D cases. The influence of the high birth-rate is presumably higher in 2010 than it is in later population projections. Consequently, method 1) and 2) are apparently less strongly affected thereof. Nonetheless, the age groups above 85 years comprise only few people. Therefore, and as supported by Fig 3 in the main text, the issue is irrelevant regarding the absolute number of future T2D cases.

To validate the prevalence projections, we refer to Tönnies et al. [4] who used the same PDE and input data to project the age- and sex-specific T2D prevalence in Germany. They compare their estimates with the observed T2D prevalence in 2015 from outpatient care claims data (n = approx. 70 million) reported by Goffrier et al. [5]. For all ages below 85 years, the values of the constant prevalence of method 1) and the projected prevalence using method 2) are slightly lower than the actual prevalence in 2015. Though, the underestimation is larger when assuming constant prevalence as is done by method 1). Therefore, we infer that compared to assuming constant prevalence as in method 1), the PDE of method 2) yields values that are likely to be closer to reality.

The impact of using different population projections of the FSO on the prevalence projection seems negligible. The results for method 1), 2) and 3) across the variants show no meaningful differences (see S2 Fig).

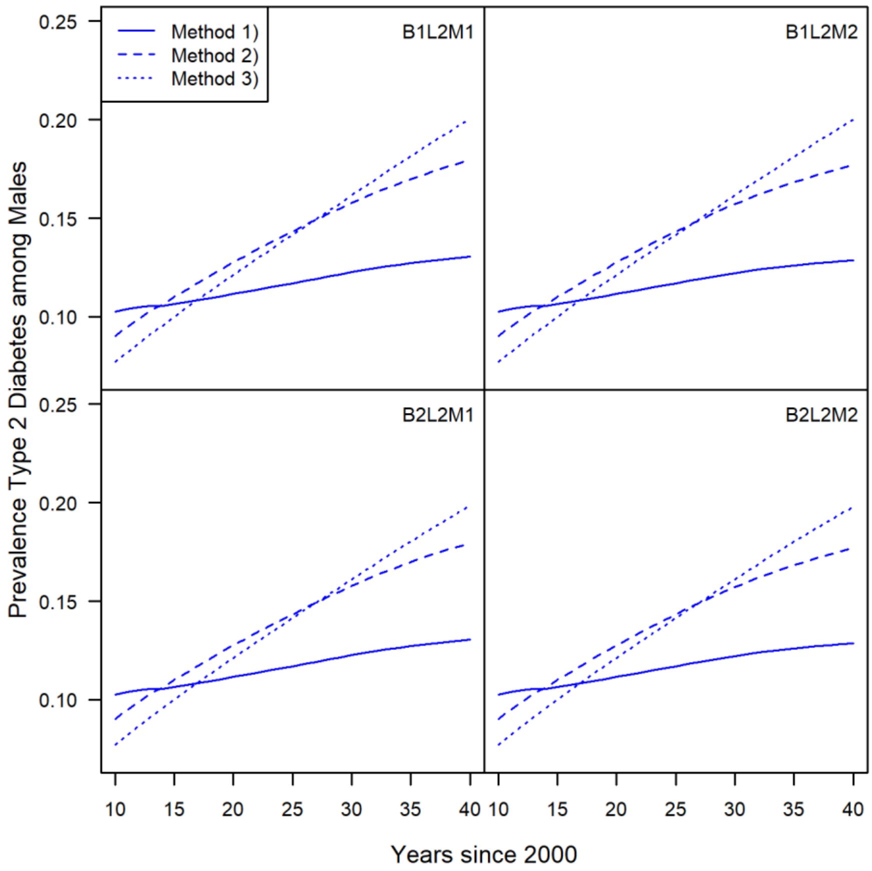


***S2 Fig. Projected prevalence across future population variants.*** *Projected overall prevalence of T2D between 2015 and 2040 in Germany among men for four population projections of the German FSO. Method 1) is based on constant age-specific prevalence; method 2) and 3) assume constant incidence rates and a 2%-decrease in the mortality rate ratio per year. The figure shows similar outcomes for the same method across the different population variants. Though, within the variants, the method's results visibly differ. We infer that the impact of the projection method on the prevalence is larger than the influence of different population projections.*

### Assessing hypothetical trends of the MRR and IR

In order to cope with potential vulnerabilities due to developments in the disease-specific environment, we reflect on 14 different scenarios with alternating the MRR and the IR. The annual changes of the age-specific IR and of the MRR are in line with those of Tönnies et al. [4]. Based on studies for Germany or countries with a similar T2D situation, we can only speculate which scenario seems most suitable: Assuming improvements in the diabetes treatment, a decreasing MRR appears plausible, while previous analyses indicate that decreasing incidence rates are more likely [4, 12]. We assumed strong incidence trends of −5% and +3% and changes in the MRR of -2% per year. S3 Fig displays the resulting case numbers for males in Germany between 2010 and 2040 using method 2) for all the considered scenarios as grey dashed lines. Applying the same scenarios to the data using method 3) yields similar results for each scenario. For a better overview and more clarity, the estimates of method 3) are thus not displayed in S3 Fig. For comparison, the results from method 1), 2) and 3) using the assumptions of our main analysis are illustrated as solid, dashed, and dotted blue line, respectively. The different scenarios cause considerably large deviations in the number of future cases. The most extreme cases project a decrease of approx. 0.3 million (-11%; stable MRR & IR -5%) or an increase of 6.4 million (216%; MRR -2% & IR +3%) male T2D cases between 2010 and 2040 for Germany (S3 Fig). The results show how susceptible estimated future case numbers are to changes in disease-specific factors. Consequently, these aspects should be central components of projection methods.

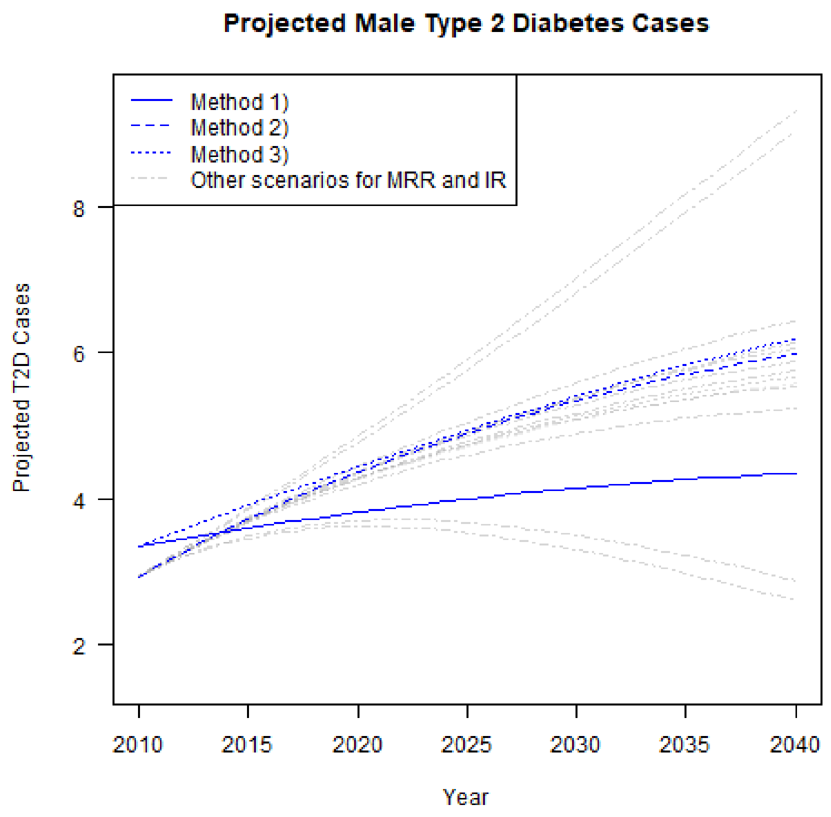


*__S3 Fig. Projected male T2D cases using different scenarios for the MRR and IR.__ The 14 different scenarios (grey dashed lines) of the MRR and IR implemented in method 2) cause considerably large deviations in the number of future cases. Some scenarios are rather extreme cases and thus, less likely. Though, it is noteworthy here how susceptible results are to changes in disease-specific input. Blue solid line represents the results of method 1) and assuming constant prevalence. Blue dashed line and blue dotted line represent method 2) and 3) assuming a constant IR and a decrease of 2% in the MRR, respectively.*

### Assessing uncertainty due to sampling error in a Monte Carlo simulation

To cope with sampling error in the input values, we applied a Monte Carlo simulation. By randomly drawing input values for the T2D IR, the MRR and the prevalence in 2010, i.e., our input year. Based on the standard error of the original estimate in 2010, we created a normally distributed error which we then added to the original estimate.

S4 Fig shows the empirical median of projected male T2D cases using variant B1L2M1 with 5000 bootstraps for method 1), 2) and 3). Again, we assume constant IR and a decreasing MRR by 2% per year for these methods. Based on this, we calculated the 2.5% and 97.5% quantiles as bounds of the 95% confidence intervals (CI) for each method. The intervals provide the range of plausible values and thus, indicate the preciseness of our future T2D case estimates.

The empirical median of the projected number of male T2D cases in Germany in 2040 amounts to 6.0 million for both methods 2) and 3). The minima and maxima of the estimated future cases in 2040 using the Monte Carlo approach range from 5.3 to 6.1 for method 2) and from 5.8 to 6.2 for method 3). The CIs overlap to a great extent which allows to conclude that there is no evidence for a statistically significant difference in the median values. Most striking is the relatively narrow range of the CIs for both methods in contrast to the large deviations of future T2D cases caused by different projection scenarios of the MRR and IR. Results may deviate from the median of future male T2D cases in Germany in 2040 by approximately 7.2% and 3.5% for method 2) and 3), respectively. In other words, the uncertainty due to sampling error in the input values is relatively small compared to the uncertainty evoked through the unknown future development of the MRR and IR.

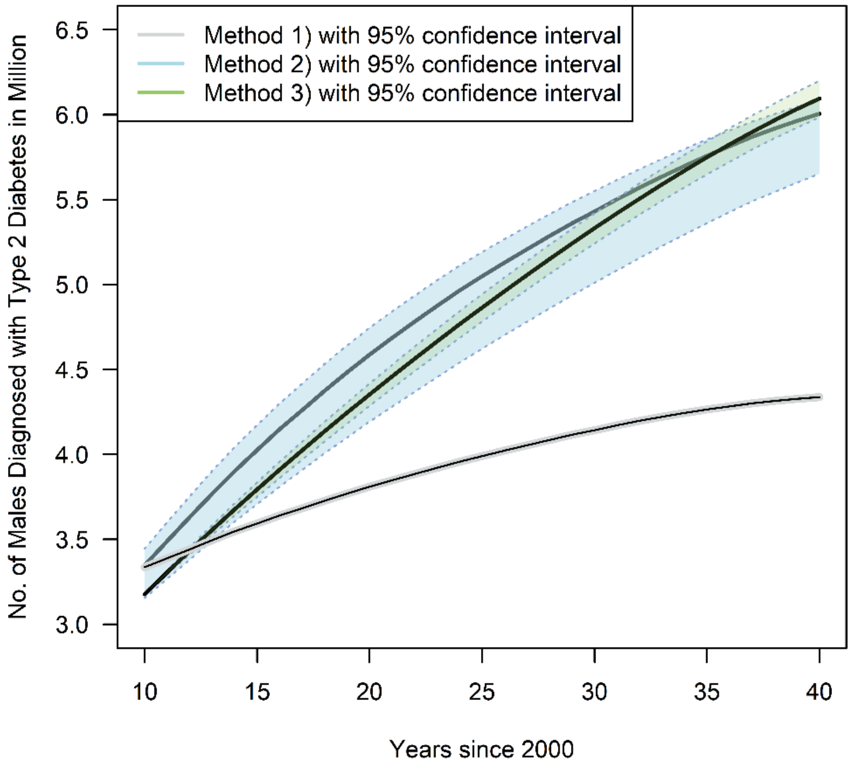


***S4 Fig. Projected number of T2D cases in Germany among men between 2015 and 2040.*** *The light-grey shaded areas show the upper and lower 95% confidence bounds for method 1). Light-blue dashed lines show the results from our Monte Carlo simulation using method 2) and assuming constant T2D*

*incidence rate and a decreasing MRR. The shaded areas in green represent the confidence intervals for method 3) and assuming constant T2D incidence rate and a decreasing MRR.*
