## Supplementary material for "Future prevalence of type 2 diabetes – a comparative analysis of chronic disease projection methods": S1 Appendix: R-Code

03/11/2021

#### Introduction

We apply the theoretical methods developed in our paper in the context of Type 2 Diabetes (T2D) to project male case numbers for Germany between 2010 and 2040.

The final paper can be found at: XXX

The R project in this file provides information about and the code for the data pre-processing and main analysis.

#### General Setup

We start with a general setup of the working environment and load the initially needed openly available packages and self-written functions.

```
rm(list=ls(all=TRUE))
setwd() # set accordingly

# Load required packages
library(deSolve)

# Load required self-written functions
# (could be outsourced and loaded via source())

# mort_L2() extrapolates the general mortality of the population
# prognosed mortality rate based on KoBev 14, calculation model 7 (M7, G2-L2-W0)
mort_L2 <- function(t, a, isMale_  = TRUE)
  {
   # year_ = 0 corresponds to 2019 (begin of 14th FSO population projection)
   year_      <- t - 19 # extrapolate m to years before 2019
   age_       <- a
   intercept_ <-  -11.431896 - 0.010102   * year_
   slope_     <-  1.086e-01 - 4.918e-06 * year_
   return(exp(intercept_ + age_ * slope_))
}

# fct_i() projects the incidence rate to the future
fct_i <- function(t, a, annInc = 1.0, isMale = TRUE){
```

```
  # here: t in years past 2000, i.e. t = 10 is 2010
  # annInc, i.e., annual increase, is yearly factor for incidence increase
  # annInc = 1.01 means 1% increase per year
  return(exp((t-10)*log(annInc))*pmax(fct_i.m(a), 0))
}

# fct_MRR() uses the MRR data from Schmidt et al. (2021) to estimate
# p(m1-m0) = (p(R-1)*m)/(p(R-1)+1) in rhs.R
fct_MRR <- function(t, a, annInc = 1.0, isMale = TRUE){
   # annInc, i.e., annual increase, is yearly factor for incidence increase
  thisR  <- c(6.87,5.01,3.79,3.27,2.46,2.16,1.85,1.67,1.71,1.65,1.45,1.29,1.20,
              1.11) # MRR for known diabetes (Schmidt et al. 2021)
  thisA  <- c(32.5, 37.5, 42.5, 47.5, 52.5, 57.5, 62.5, 67.5, 72.5, 77.5, 82.5,
              87.5, 92.5, 97.5) # corresponding age groups
  retval <- pmax(1.11, exp((t-10)*log(annInc) + approx(thisA, log(thisR), a, rule=2)$y)
                 ) # 1.11 is minimum MRR
  return(retval)
}

# define initial (t = 2010) age-specific prevalences from Tamayo et al. (2016)
fct_p0 <- function(a, isMale = TRUE) {
     return(pmax(fct_p0.m(a), 0))
}

# init_splines() interpolates the initial values for the age- and sex-specific
# prevalence and incidence
init_splines <- function(){
   # prevalence in base year (t=0)
   as.p0    <-  c(17.5+5*(0:16), 100)
   p0       <-  c(0.06,0.11,0.19,0.44,1.12,2.17,4.03,7.28,11.87,17.03,20.32,
                  23.45,26.36,26.27,24.58,23.66,19.69,16.52) # in %
   fct_p0.m <<- splinefun(as.p0, 0.01*p0, method="nat")

   # incidence
   as.inc   <-  22.5+5*(0:15)
   inc_     <-  c(0.21,0.49,1.18,2.48,4.58,7.64,11.54,15.83,19.78,23.13,25.77,
                  27.74,28.12,26.71,24.69,21.21)
   fct_i.m  <<- splinefun(as.inc, 0.001*inc_, method="nat")
}

# call the function
init_splines()
```

Second, we load the required data. Being more precise, here we need input values on the future population in Germany. For our present application, we use the data of the 14th coordinated population projection of the Federal Statistical Office (FSO, in German "Destatis") released in 2018 and which covers the period up to 2060. It contains several variants and model calculations which illustrate the range of possible developments influenced by the demographic components of fertility, mortality and migration. We restrict our analyses to the time horizon until 2040, to the male population and one of the variants of the FSO (variant G1L2W1).

```
# Read the data from the FSO variants containing the future population numbers
# For simplicity, we use only variant G1L2W1 for males in this demo
file <- c("./data/G1L2W1_males.txt")
```

```
g1l2w1_m <- read.csv(paste(file, sep=""), header=TRUE, sep=";")

# Restructure and collect the population scenario data in one array
#                            scenario  years      age
N_m <- array(data = NA, dim = c(  1,     31,      101))
N_m[1,,] <- as.matrix(g1l2w1_m[1:31, 2:102])


# Set hard-coded variables (explained in the following, may change later)
ScRi  <- c(1:14) # 14 scenarios for MRR and IR (DdzSz)
ScFSO  <- 1   # 1 scenarios from FSO population projections (baSz)
myages <- 18:100 # relevant ages, 18 means 18-19-years-old, coded as myages+1
```

### Main Analysis

#### Method 2)

##### Prevalence Projection using Method 2)

In a first step, we model the prevalence of T2D in Germany for men until 2040 considering 14 different scenarios. Therefore, we use a partial differential equation (PDE) as suggested by Tönnies et al. (2019). The scenarios reflect temporal trends in the mortality rate ratio (MRR) and the incidence rate (IR, in Code called i or i_). The MRR is defined as the ratio of the mortality rates of people with vs. without diagnosed T2D. The mortality rate of people with T2D is higher compared to the mortality of people without the disease, hence, we incorporate the MRR into the equation to reflect on this issue.

```
# Set up the equation of the PDE to compute dp_ (i.e., change in prevalence)
rhs <- function(a, p, parms) {
  with(as.list(parms),
       {
         MRR_  <- fct_MRR(t0 + a, a, annInc = rFac, isMale = isMale) # MRR
         i_    <- fct_i(t0 + a, a, annInc = iFac, isMale = isMale) # IR
         m_    <- mort (t0 + a, a, isMale = isMale) # general mortality
         dp_   <- (1-p)*(i_ - m_ * p * (R_ - 1)/(p * (R_ - 1) +1)) # PDE
         list(dp_)
       }
  )
}

# Set up array to collect the respective prevalence (p_ml2)
# Data for males with mortality based on FSO scenarios with life expectancy L2
#                                  ages 0-100, years 2010-40, MRR & IR scenarios
p_ml2 <- array(data = NA, dim = c(        101,            31, 14))

# Set up matrix to hold possible temporal trends for the MRR and the IR
Sc_Ri <- matrix(c(1:14, 1.00,  1.00,  1.00,  1.00,  1.00, 1.00, 1.00, 0.98,
                  0.98,  0.98,  0.98,  0.98, 0.98, 0.98,
                  1.00, 0.999, 0.995, 1.001, 1.005, 1.03, 0.95, 1.00, 0.999,
                  0.995, 1.001, 1.005, 1.03, 0.95),
                nrow = 14, ncol = 3,
                dimnames = list(1:14, c("Sc", "R", "i")))
```

Having setup the mathematical model, we loop over the years and the ages (one could add the sex, but for simplicity, we restrict the analysis for males in Germany) to compute the age- and sex-specific prevalence.

```
# first, loop over all ages for each scenario of MRR and IR and compute the
# age-stratified prevalence
for(Sc in ScRi) # scenarios for the MRR and IR
{
  for(as in 0:100)
  {
    y_         <- fct_p0(as, TRUE)
    a_         <- seq(as,(as+30), by = 1)
    x          <- c(p = y_)
    rf_        <- Sc_Ri[Sc, 2] # 2% decrease in the MRR
    if_        <- Sc_Ri[Sc, 3] # IR constant
    parms      <- c(isMale = TRUE, mort = mort_L2, t0 = 10 - as,
                    rFac = rf_, iFac = if_) # t0 in years after 2000
    out_       <- as.data.frame(rk4(x, a_, rhs, parms))
    for(ii in 1:min(31, nrow(p_ml2) - as)){
      # using nrow(p_m) to avoid that subscripts are out of bound
      p_ml2[as + ii, ii, Sc] <- out_$p[ii]
    }
  }
}

# loop over all years of overall time horizon to project the prevalence to 2040
for(Sc in ScRi) # scenarios for the MRR and IR
{
  for(ts in 11:40) # from year 2010 until year 2040
  {
    y_         <- 0
    a_         <- seq(0, (40-ts), by=1)
    x          <- c(p = y_)
    rf_        <- Sc_Ri[Sc, 2]
    if_        <- Sc_Ri[Sc, 3]
    parms      <- c(isMale = TRUE, mort = mort_L2, t0 = ts,
                    rFac = rf_, iFac = if_)
    out_       <- as.data.frame(rk4(x, a_, rhs, parms))
    for(ii in 1:(40-ts+1)){
      p_ml2[ii, ts - 10 + ii, Sc] <- out_$p[ii]
    }
  }
}
```

##### Case Projection using Method 2)

Since concrete case numbers rather than relative prevalence are of interest, we apply the projected T2D prevalence to the population projections of the FSO. We receive results for case numbers and also the number of non-diseased people with regards to the disease of interest.

```
# create arrays to collect the resulting total case/ non-case numbers per year
                     #FSO variants, years since 2010, MRR/IR scenarios
C_m  <- array(data = NA, dim = c(1,            31,      16)) # for cases
NC_m <- array(data = NA, dim = c(1,            31,      16)) # for non-diseased
```

```r
# apply projected prevalence to future population figures
for(Sc in ScRi) # scenarios for the MRR and IR
{
  for(FSOvar in ScFSO) # FSO variants of population projection
  {
    for(year in 1:31) # year = 1 equals year 2010
    {
      C_m[FSOvar, year, Sc] <-
        sum(p_ml2[myages+1, year, Sc]*N_m[szNr, year, myages+1], na.rm = T)
      # -> p_m[18+1,] equals 18-19-years old
      NC_m[FSOvar, year, Sc] <- sum(N_m[FSOvar, year, myages+1], na.rm = T)
    }
  }
}
```

#### Method 1)

##### Case Projection using Method 1)

With method 1), we use the initial age-specific prevalence as is in 2010 (Tamayo et al., 2016). This method assumes that the prevalence remains constant over the whole time horizon. The constant prevalence is simply multiplied with population projections without reflecting on any disease-specific dynamics.

```r
# we use scenario 15 to save results for method 1)
Sc <- 15

for(FSOvar in ScFSO) # FSO variants
{
  for(year in 1:31) # year = 1 equals year 2010
  {
    C_m[FSOvar, year, Sz] <-
      sum(p_ml2[myages+1, 1, 1]*N_m[FSOvar, year, myages+1], na.rm = T)
    NC_m[FSOvar, year, Sz] <- sum(N_m[FSOvar, year, myages+1], na.rm = T)
  }
}
```

#### Method 3)

##### Case Projection using Method 3)

With method 3) we use two PDEs to directly compute changes in the number of cases and the number of non-diseased people with regards to T2D. Thus, in this case we do not need to project the prevalence in a first step. This method accounts for underlying changes in disease-specific rates (i.e., MRR and IR). Further, we

```r
# create arrays for resulting male age-specific cases & numbers of non-diseased
#                                  ages, 2010-40
myC_m <- array(data = NA, dim = c(101,     31))
myS_m <- array(data = NA, dim = c(101,     31))

pp_m <- p_ml2[myages+1,1,8] # use prevalence from 2010 to split general
```

```r
# mortality into m0 and m1, i.e., mortality rate of non-diseased and diseased

fct_m0       <- function(t, a, annInc = 1.0, isMale){
                   thisP <- pp_m
                   pp_     <- approx(myages, thisP , xout = a, rule = 2)$y
                            # linearly interpolate between given data points
                            # (here prevalence data)
                   RR_     <- fct_MRR(t, a, annInc = 1.0, isMale)
                   mm_     <- mort_L2(t, a, isMale)
                   return(annInc^(t-19)*mm_/(1 + pp_*(RR_ - 1)))
                }
fct_m1       <- function(t, a, annInc = 1.0, isMale){
                   return(fct_MRR(t, a, annInc = 1.0,  isMale) *
                            fct_m0(t, a, 1.0, isMale) * annInc^(t-15))
                }

# set up the PDEs used with method 3)
rhsSC <- function(a, SC, parms) {
  with(as.list(c(SC, parms)),
       {
         m0_      <- fct_m0(t0 + a, a, annInc = 0.995, isMale = isMale)
         m1_      <- fct_m1(t0 + a, a, annInc = 0.975, isMale = isMale)
         i_       <- fct_i (t0 + a, a, annInc = 1.000, isMale = isMale)
         dS_      <- -S*(i_ + m0_) # change in the number of non-diseased
         dC_      <- i_ * S - m1_ * C # change in the number of cases
         list(c(dS_, dC_))
       }
  )
}

FSOvar <- 1 # using variant G1L2W1 from the FSO population variants

# loop over all ages and then all years incl. in the projection
for(as in 0:100){
   p_       <- ifelse(as > 18, pp_m[as - 18 + 1], 0)
   y0_      <- (1-p_)*N_m[FSOvar, 2, as+1]
   y1_      <-     p_*N_m[FSOvar, 2, as+1]
   a_         <- seq(as,(as+30), by = 1)
   x          <- c(S = y0_, C = y1_)
   parms      <- c(isMale = TRUE, t0 = 10 - as) # t0 in years after 2000
   out_       <- as.data.frame(rk4(x, a_, rhsSC, parms))

   for(ii in 1:min(31, nrow(myC_m) - as)){
     # nrow(p_m) to avoid that subscripts are out of bound
     # mat[age + future years, years]
     myS_m[as + ii, ii] <- out_$S[ii]
     myC_m[as + ii, ii] <- out_$C[ii]
   }
}

for(ts in 11:40){
   a_         <- seq(0, (40-ts), by=1)
   x          <- c(S = N_m[baSz, ts - 10, 1], C = 0)
```

```r
  parms      <- c(isMale = TRUE, t0 = ts)
  out_       <- as.data.frame(rk4(x, a_, rhsSC, parms))
  for(ii in 1:(40-ts+1)){
    myS_m[ii, ts - 10 + ii] <- out_$S[ii]
    myC_m[ii, ts - 10 + ii] <- out_$C[ii]
  }
}
```
